# Prevalence of SARS-CoV-2 infection among COVID-19 RT-PCR laboratory workers in Bangladesh

**DOI:** 10.1101/2021.12.02.21267191

**Authors:** Mohammad Jahidur Rahman Khan, Samshad Jahan Shumu, Ruksana Raihan, Nusrat Mannan, Md. Selim Reza, Nazia Hasan Khan, Amirul Huda Bhuiyan, Paroma Deb, Farzana Mim, Arifa Akram

**Affiliations:** Department of Microbiology, Shaheed Suhrawardy Medical College, Dhaka, Bangladesh; Department of Microbiology, US-Bangla Medical College, Dhaka, Bangladesh; RT-PCR LAB, Bangabandhu Sheikh Mujib Medical College, Faridpur, Bangladesh; RT-PCR LAB, United Hospital Limited, Dhaka, Bangladesh; RT-PCR LAB, Narayanganj 300 Bed Hospital, Narayanganj Bangladesh; Department of Virology, Dhaka Medical College, Dhaka, Bangladesh; Department of Biochemistry & Molecular Biology, Jahangirnagar University, Dhaka, Bangladesh; Department of Virology, National Institute of Laboratory Medicine and Referral Centre, Dhaka, Bangladesh

**Keywords:** RT-PCR laboratory, Quality control, Personal protection equipment, Risk factor Laboratory health worker, Medical technicians, Contamination

## Abstract

**Background:** Health care workers (HCWs) at the frontline are confronting a substantial risk of infection during the coronavirus disease 2019 (COVID-19) pandemic. This emerging virus created specific hazards to researchers and laboratory staff in a clinical setting, underlined by rapid and extensive worldwide transmission. This study aimed to investigate the prevalence of SARS-CoV-2 infection among COVID-19 RT-PCR laboratory health workers in Bangladesh.

**Materials & Methods:** This retrospective study was conducted between October 2 to December 2, 2020. A total of 508 participants, including doctors, scientific officers, medical technologists, and cleaners working in several COVID-19 RT-PCR laboratories, were included in this study. Data were collected from each participant using a semi-structured questionnaire prepared in the format of an anonymous Google form. All participants provided informed consent. The Ethical clearance was obtained from the Institutional Ethics Review Committee of Shaheed Suhrawardy Medical College, Dhaka, Bangladesh. All statistical analyses were performed using SPSS (Statistical Package for the Social Sciences) version 25.0 software (SPSS, Inc).

**Results:** Out of the 508 participants, 295 tested positive for SARS CoV-2 RT-PCR. Among the positive cases, 202 were men, 93 were women, with the median age of 30 years. The most positive cases were medical technologists (53.22%) followed by doctors (28.8%). Out of the 271 symptomatic positive cases, the most typical symptoms were fever (78.5%), fatigue (70%), loss of smell and taste (65%), cough (64%), and others. Hypertension, obesity, and diabetes were found in 8.8%, 8.8%, and 7.1% positive cases. A + blood group was present in 37% of the positive cases, followed by the B+ blood group (27%) and O+ blood group (25%). Inadequate supply of personal protection equipment (PPE), absence of negative pressure ventilation, laboratory contamination, and no training on molecular test methods were found in 13.8%, 67.8%, 44.7%, and 40.6% of positive cases, respectively.

**Conclusion:** Evaluating the infection status of laboratory health workers is crucial for drawing attention from the public, providing practical suggestions for government agencies, and increasing protective measures for laboratory health workers.

## Introduction

Since its discovery, SARS-CoV-2 has shaken out of the world and has become a pandemic. As of September 4, 2021, there were 220,362,472 reported cases and 4,562,679 deaths worldwide^1^. In Bangladesh, the first case of SARS-CoV-2 infection was confirmed on March 8, 2020. Subsequently, Bangladesh was facing an increasing risk of imports and some local cluster cases of COVID-19. As of September 4, 2021, there are 1,510,283 confirmed cases and 26,432 deaths in Bangladesh^2^.

Health care personnel around the globe have the most significant risk of getting infected and infecting others in their surrounding environment^3^. According to initial estimation, healthcare workers account for 10%–20% of all confirmed cases^4^. During the pandemic, medical services worldwide is facing an unavoidable burden of public health challenges^5^. In Bangladesh, most molecular laboratories performed RT-PCR to detect SARS-CoV-2 have been established after the pandemic began. These facilities were confronted an increased amount of real-time reverse transcriptase-polymerase chain reaction (RT-PCR) testing of SARS-CoV-2 for patients suspected as Covid-19, quarantined healthcare workers; travelers came back from high-risk countries as well as other required samples. The staff available for the laboratory was swiftly deployed to receive the large number of clinical samples without adequate amounts of training and PPEs. To confront this novel coronavirus never experienced before, some public health laboratory workers overlooked concerns about the possible risks of SARS-CoV-2 infection from their occupational exposure. While the protection of laboratory healthcare workers during the COVID-19 pandemic is one of the primary concerns, data regarding this issue are still inadequate^6^.

At present, around 1200 health workers, including doctors, microbiologists, biochemists, molecular biologists, medical technologists, cleaners, are working in over 100 COVID-19 RT-PCR laboratories across the country^7^. Many of them were infected by SARS-CoV-2 during this ongoing pandemic. They become a source of contamination in many laboratories^8^. The testing capacity of a COVID-19 RT-PCR laboratory is reduced when several workers become SARS-CoV-2 infected. The physical environment of the laboratory and workload play an essential role in transmitting SARS-CoV-2 among the laboratory workers^9^. The chance of getting infected by SARS-CoV-2 also depends on a laboratory health worker’s age, comorbidity, and functional skill^10^.

Thus, we conducted a retrospective study to investigate the prevalence of SARS-CoV-2 infection among COVID-19 RT-PCR laboratory health workers in Bangladesh and assess the underlying factors related to the high infection rate of SARS-CoV-2.

## Methods

### Study Design and data collection

We conducted a retrospective online survey from October 2 to December 2, 2020. A semi-structured questionnaire was prepared using an anonymous Google form. The generated link was shared with the focal persons of each laboratory and several Facebook and WhatsApp groups involving doctors and medical technologists. We decided to collect the data using online approaches and maintain social distance during Bangladesh’s pandemic condition. Additional data were collected from some participants who did not fill the Google form entirely over the telephone. A hard copy of the questionnaire was also supplied to some participants who were not habituated with online submission by Google form.

### Study population

Total 534 laboratory health workers, including doctors, scientific officers (microbiologist, biochemist, molecular biologist), medical technologists, and cleaners, filled up the Google form. Twenty-six participants were excluded as they had COVID-19 like symptoms, but RT-PCR did not confirm the diagnosis. The remaining 508 laboratory health workers from multiple COVID-19 RT-PCR laboratories were included in this study. Informed consent was obtained from all participants.

### Statistical Analysis

The confirmed COVID-19 cases among HCWs were categorized according to the following parameters: sex, occupation type, hospital type, infection status, and others. All statistical analyses were performed using SPSS (Statistical Package for the Social Sciences) version 25.0 software (SPSS, Inc).

## Results

Among the 508 participants, 295 (58%) were tested positive for SARS-CoV-2 RT-PCR, and 237 (80.3%) were between the 24-44 years age group; male participants were 68.5%, and females were 31.5% (Table 1). Most participants were medical technologists (53.7%), followed by doctors (27.2%) (Table 2). Among the 295 positive cases, 271 were symptomatic. Analyzing the symptoms, we found 78.5% of them had fever, fatigue (70%), loss of smell and taste (65%), cough (64%), and others (Figure 1). Among the positive cases, the A+ blood group (37%) was affected more by COVID-19, followed by the B+ blood group (27%) (Figure 2). We analyzed their comorbidity status and found that hypertension and obesity were most common, 8% in both cases, followed by diabetes (7%) (Table 3). Among the positive cases, 13.8% had not an adequate supply of personal protection equipment (PPE), 67.8% had not the negative pressure ventilation system, 44.7% had an incidence of laboratory contamination, and 40.6% did not get any kind of training on molecular test methods or quality control (QC) (Table 4).

**Table 1:** Demographic data of the study population.

|  |  | SARS-CoV-2 RT-PCR |  |
| --- | --- | --- | --- |
|  | Total | Positive | Negative |
|  | 508 (100%) | 295 (58%) | 213 (42%) |
| Age (years) |  |  |  |
| Median | 30 | 30 | 30 |
| <24 (%) | 50 (9.8%) | 27 (9.2%) | 23 (10.8%) |
| 24-44 (%) | 413 (81.3%) | 237 (80.3%) | 176 (82.6%) |
| >44 (%) | 45 (8.9%) | 31 (10.5%) | 14 (6.6%) |
| Sex |  |  |  |
| Male (%) | 344 (67.7%) | 202 (68.5%) | 142 (66.7%) |
| Female (%) | 164 (32.3%) | 93 (31.5%) | 71 (33.3%) |

**Table 2:** Infection rate according to the designation of laboratory workers.

| Designation | Total participants<br>(n= 508) | SARS-CoV-2 RT-PCR |  |
| --- | --- | --- | --- |
|  |  | Positive (n= 295) | Negative (n= 213) |
| Doctor | 138 (27.2%) | 85 (61.6%) | 53 (38.4%) |
| Scientific officer | 53 (10.5%) | 27 (50.9%) | 26 (49.1%) |
| Medical Technologist | 273 (53.7%) | 157 (57.5%) | 116 (42.5%) |
| Cleaner | 44 (8.6%) | 26 (59.0%) | 18 (41.0%) |

**Table 3:** Comorbidities found in SARS-CoV-2 RT-PCR-positive and SARS-CoV-2 RT-PCR-negative cases.

| Comorbidity | Total participants<br>(n= 508) | SARS-CoV-2 RT-PCR |  |
| --- | --- | --- | --- |
|  |  | Positive (n= 295) | Negative (n= 213) |
| DM | 28 (5.5%) | 21 (7.1%) | 07 (3.3%) |
| HTN | 38 (7.5%) | 26 (8.8%) | 12 (5.6%) |
| Asthma | 21 (4.1%) | 15 (5.0%) | 06 (2.8%) |
| Obesity | 44 (8.6%) | 26 (8.8%) | 18 (8.4%) |
| IHD | 06 (1.1%) | 05 (1.7%) | 01 (0.5%) |

**Table 4:** Association of risk factors among SARS-CoV-2 RT-PCR-positive cases.

| **Risk factors** | **SARS-CoV-2 RT-PCR-positive cases** |
| --- | --- |
| Inadequate supply of standard PPE | 13.8% |
| Absence of negative pressure ventilation | 67.8% |
| Incidence of laboratory contamination | 44.7% |
| No training on molecular test methods | 40.6% |

**Figure 1:**
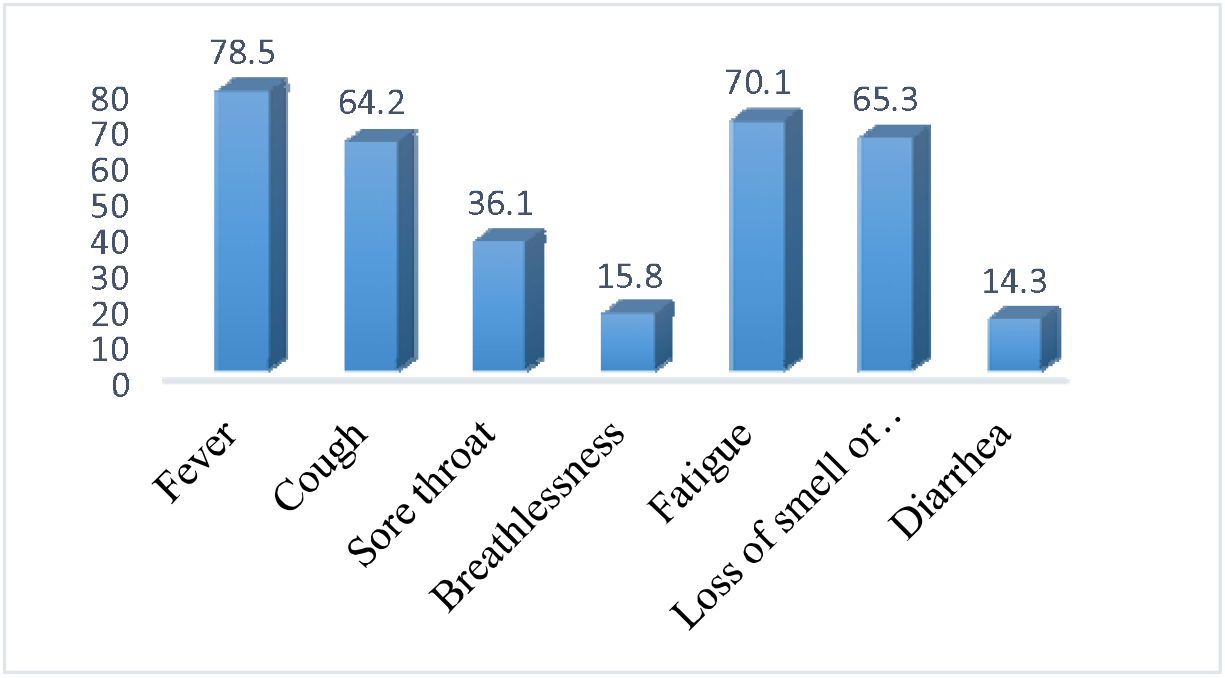
Symptoms of the SARS-CoV-2 RT-PCR-positive cases (%)

**Figure 2:**
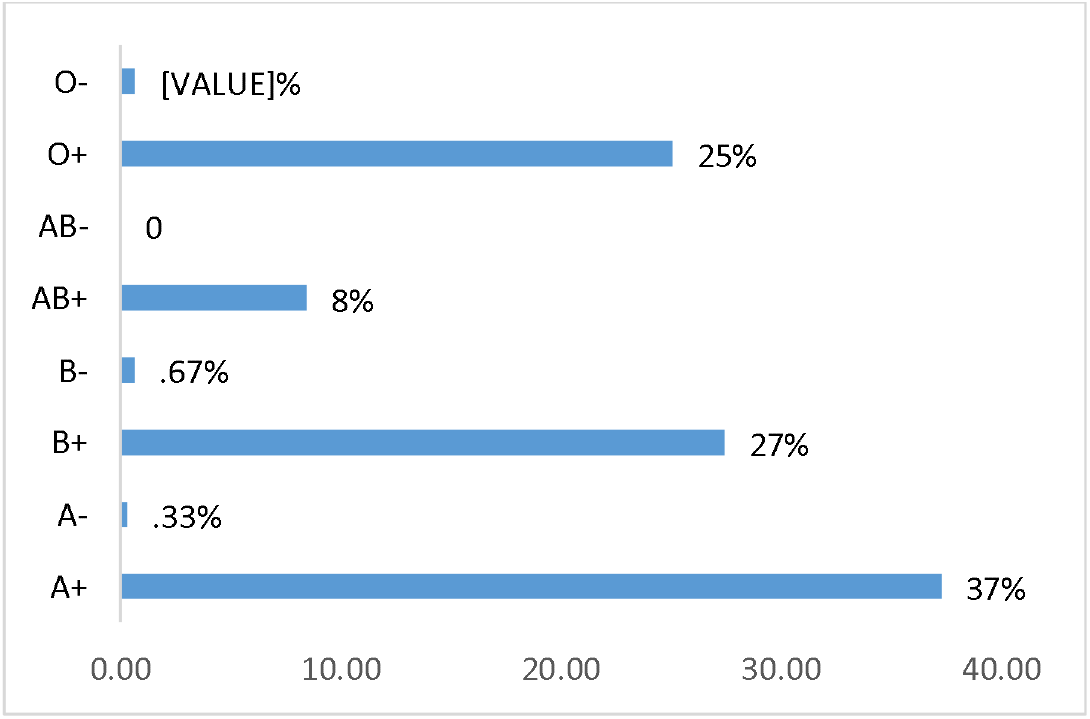
Blood group distribution among the SARS-CoV-2 RT-PCR-positive cases.

## Discussion

Findings from a previous pandemic of other coronaviruses revealed that frontline healthcare workers (HCWs) were at the highest risk of infection because of close contact with infected patients, touching the contaminated surfaces, the hiding of epidemiological histories by patients, inadequate training for infection prevention and control and conducting the high-risk procedures in airway management^11,12^. Additional laboratory professionals, including virologists, microbiologists, medical technologists, cleaners, are also at high risk through exposure to specimens collected from SARS-CoV-2 infected patients. This study retrospectively collected epidemiological and related data of laboratory personnel working in multiple COVID-19 RT-PCR laboratories. Among the 508 participants of our research, we found 295 (58%) lab workers became positive during their services, and most of them were male and young (24-44 years age group).

Among laboratory health workers, medical technicians (MTs) possess a higher risk of regular handling of both symptomatic and asymptomatic cases^13^. Our study also found that medical technicians affected more almost 53% of cases. Analyzing the symptoms of positive cases, we found fever (78%), fatigue (70%), loss of smell and taste (65%), cough (64%), breathlessness (15%), and diarrhea (14%). Most of the cases were symptomatic (91%). A meta-analysis study on COVID-19 comorbidities shows that the most common comorbidities identified are hypertension (15.8%), which also matched our research; we found it in 8% of cases^14^. Blood group A had a significantly higher risk for acquiring COVID-19 than other blood groups in our study, which is also matched with the study of Barcelona^15^.

SARS-CoV-2 can be transmitted during the incubation period when a patient has nonspecific symptoms or no symptoms at all^14^. Therefore, it is necessary to protect them from SARS-CoV-2 infection, and additional transmission-based precautions should be taken^15^. HCWs infected by SARS-CoV-2 can increase the risk of transmission, and their absence from work can decrease health service performance. These may disrupt the chain management of a transmission^16^. To minimize the risk of transmission, healthcare workers should be provided with sufficient personal protective equipment (PPE) supplies, training on infection control, maintenance of personal hygiene, waste management^17^. Laboratory staff is advised to use PPE like the surgical or N95 mask, gowns, shield in the correct order, and they must be trained about it. Several studies have suggested that factors such as sufficient supplies of PPE, hands-on training on how to use them, etc., perform a crucial role in controlling such infections, which notably decreases the risk of transmission^18^. During the pandemic, especially at the initial stage, the global scarcity of masks, respirators, face shields, and gowns developed due to the sudden increase in demand and supply chain interference. Therefore, laboratory workers must preserve PPE by increased use or reuse, and infection prevention and control protocols could be maintained for the same reason^19^. Our study revealed that 13.8% of SARS-CoV-2 infected laboratory workers had an inadequate supply of PPE, and 67.8% had no negative pressure ventilation system in their workplace. 40.6% of SARS-CoV-2 infected laboratory workers did not train on molecular test methods or quality control (QC).

Healthcare workers play an essential role in in-hospital transmission. They are a potential source of nosocomial infection^20,21,22^. In severe acute respiratory syndrome coronavirus (SERS-CoV) and Middle East respiratory syndrome coronavirus (MERS-CoV) infection, nosocomial outbreaks have played a crucial part in spreading these viruses. The proportions of nosocomial conditions with early outbreaks of COVID-19, SARS, and MERS were 44.0%, 36.0%, and 56.0%, respectively^23^. In our study, 44.7% of SARS-CoV-2 infected laboratory workers gave a history of laboratory contamination within six months.

Our study has some notable limitations. First, we could not collect biochemical data from all HCWs and could not manage the duration of hospital stay. Moreover, we did not obtain Whole Genome Sequencing of the healthcare workers who tested positive for SARS-CoV-2, and analyses should be interpreted with caution because of the small sample size.

## Conclusion

The safety of laboratory health workers should be confirmed to end the pandemic, as COVID-19 is ongoing. In this study, we tried to analyze the infection status of laboratory health workers as it is not done before in Bangladesh, and it is essential to attract enough attention from the government and public. It will draw the attention of the government and non-government agencies to maintain the QC of COVID-19 RT-PCR laboratories, to improve protective measures like the adequate supply of PPE, and to arrange more hands-on training for laboratory health workers.

## Data Availability

The datasets used and analyzed during the current study are available from the corresponding author on reasonable request.

## Declarations

### Ethics approval and consent to participate

This study was approved by the Ethical Review Committee of Shaheed Suhrawardy Medical College, Dhaka, Bangladesh (No-ShSMCH/Ethical/2021/08).

### Consent for publication

Written informed consent was taken from each participant.

### Authors’ contributions

SJS, NM and RR supervised the study. NR, AHV and PD take part in conceptualization and methodology. AA and FM analyzed and interpreted the patient data. MSR performed the qRT-PCR. MJRK was the main contributor in writing the manuscript. All authors read and approved the final manuscript.

### Conflict of Interest and Authorship Conformation Form

All authors have participated in (a) conception and design, or analysis and interpretation of the data; (b) drafting the article or revising it critically for important intellectual content; and (c) approval of the final version. This manuscript has not been submitted to, nor is under review at, another journal or other publishing venue. The authors have no affiliation with any organization with a direct or indirect financial interest in the subject matter discussed in the manuscript.

## Acknowledgments

First, we would like to express our sincere gratitude to the Institute of Epidemiology, Disease Control and Research (IEDCR) for supplying us with all types of rapid antigen test kits as part of the validation procedure. We would like to thank the principal of Shaheed Suhrawardy Medical College for permitting this study in his institute. We would also like to thank all of the doctors and laboratory staff of the Department of Microbiology for their continuous support throughout the study. We are also grateful to all of the study participants who actively participated and co-operated to make it possible to complete this study. Finally, we would like to thank all the people who have supported me to complete the research work directly or indirectly.

